# Association Between BCG Policy is Significantly Confounded by Age and is Unlikely to Alter Infection or Mortality Rates

**DOI:** 10.1101/2020.04.06.20055616

**Authors:** Stefan Kirov

## Abstract

Recently a number of publications looked at the association between COVID-19 morbidity and mortality on one hand and countries’ policies with respect to BCG vaccination on the other. This connection arises from differences in the rates of infection in countries where BCG vaccination is mandatory compared to countries where mandatory vaccination no longer exists or was never implemented in the first place. In at least 2 preprint publications the authors expressed the view that the “known immunological benefits” of BCG vaccination may be behind the biological mechanism of such observation.

One study accounted for different income levels in different groups. Another study did not attempted to do so, instead exploring the differences between countries where a booster shot is given vs others where no such practice exists (finding no connection).

Both of these studies did not explore other potential confounding factors. Meanwhile the press has focused on these headlines and pushed the narrative that BCG vaccination is causally linked to infection and mortality rates. This poses a serious challenge, demonstrated by the recently initiated clinical trials on BCG vaccination within the COVID19 context.

This study shows that population age is a very significant confounding factor that explains the rates of infections much better and has a solid biology mechanism which explains this correlation. It suggests that BCG vaccination may have little or no causal link to infection rates and advises that any follow up studies should control for several confounding factors, such as population age, ethnicity, rates of certain chronic diseases, time from community spread start date, major public policy decisions and income levels.

## Introduction

BCG vaccine has been associated in multiple studies with effects beyond protection against tuberculosis, which is the original target of the intervention(1). As such, BCG vaccination has been shown to enhance the protection provided by H1N1 vaccine(2). Another study observed that prior BCG vaccination attenuates yellow fever vaccine associated viremia(3). These findings prompted two independent studies(4,5) in which a strong correlation between the presence of BCG vaccination and a reduced rate of COVID19 infections and/or death rates was observed.

These observations were also communicated by the press (6–9). While some of these publications authors contacted additional subject matter experts who cautioned that the studies are not yet peer reviewed and urged patience(7), there is danger that the observations will be over interpreted and used for policy decisions. There are currently at least 2 ongoing studies focusing on the effect of BCG vaccination in the context of COVID19 infections(10) any decisions should be taken only after conclusive evidence is presented after the end of the trials.

At the same time, some of the established risk factors for COVID19 hospitalization and death such as age(11) and BMI(12) are easy to evaluate in the same context (per country). Given the extreme urgency and the potential for serious consequences I decided to explore this matter in greater depth.

## Materials and Methods

Data on average population age per country and number of infections was collected from Wikipedia. BCG policy and income level data was obtained from one of the original studies. BCG and rubella immunization rates were obtained from WHO website(13).

Country names were cleaned and data was merged in R (Rstudio 1.1.456 on Ubuntu 16 Linux, R version 3.2.3). All scripts and data files are available from https://github.com/kirovsa/covid19-bcg.

As with one of the original studies the countries with population of less than 1M were excluded(4). For analysis of mortality rates countries with no deaths on record were excluded as these are likely to be in the very initial phase of the epidemic and will introduce significant noise.

To determine the effect of different factors I used lm function from stats package. Evaluating factors effects on infections in the presence of random effect was done with lme from nlme package. Log likelihood was tested with lrtest from lmtest package.

BCG policy and income level were coded according to one of the studies that found the association with COVID19 infection(4):

BCG Policy

1 = current universal policy

2= used to recommend, not anymore

3 = never had universal policy

2018 FY income level:

Low income (L) -1

Lower middle income (LM)-2

Upper middle income (UM)-3

High income (H)-4

## Results

Based on a significant amount of accumulated data, age is a significant factor predicting hospitalization of COVID19 patients and fatal outcome. Younger people also seem more likely to remain asymptomatic. Therefore I decided to evaluate a linear regression model that accounts for 3 factors- BCG policy, income level and median age per country.

While the model as a whole explains very well the differences in infection rates across countries, the most significant factor was income level, followed by median age. BCG policy was significant but lagged behind the other factors (Figure 1). However, BCG immunization rates was not significant in this model at alpha level at 0.05 (p=0.088). The likelihood test did find that the BCG policy had an effect (p=0.0028) compared to the full model, however this was not true for BCG vaccination rates (p=0.08). If there is a causal link between BCG vaccination and COVID19 infection rates one would expect this association to hold or even get stronger, something I did not find evidence for.

**Figure 1.**
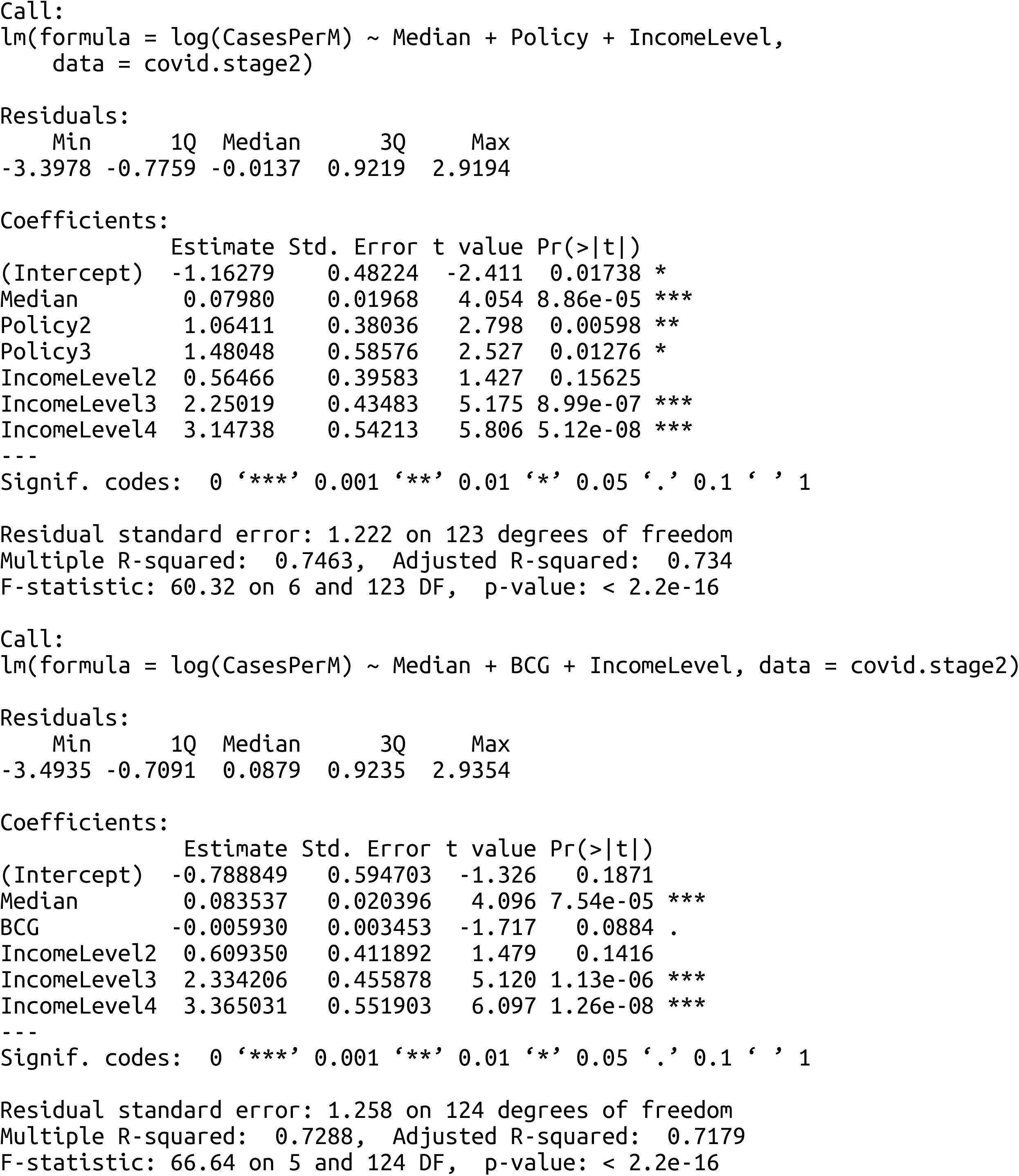
Linear regression model exploring policy, age and income level effect on infection rates

The Pearson correlation between median age and infection rates was also much higher at R=0.774 than the reported correlation between the BCG policy and the infection rates at R=0.521 or the reported correlation between start date of BCG vaccination and infection rates (R=0.21).

The correlation between number of cases per million people with the median age in a country does not change substantially between different policy categories (Figure 2A), though there was some separation between categories 1 and 3. This can only be evaluated for countries with high rates of infection and also higher median age. When the BCG immunization rates were used instead of the policy there was no association (Figure 2B).

**Figure 2.**
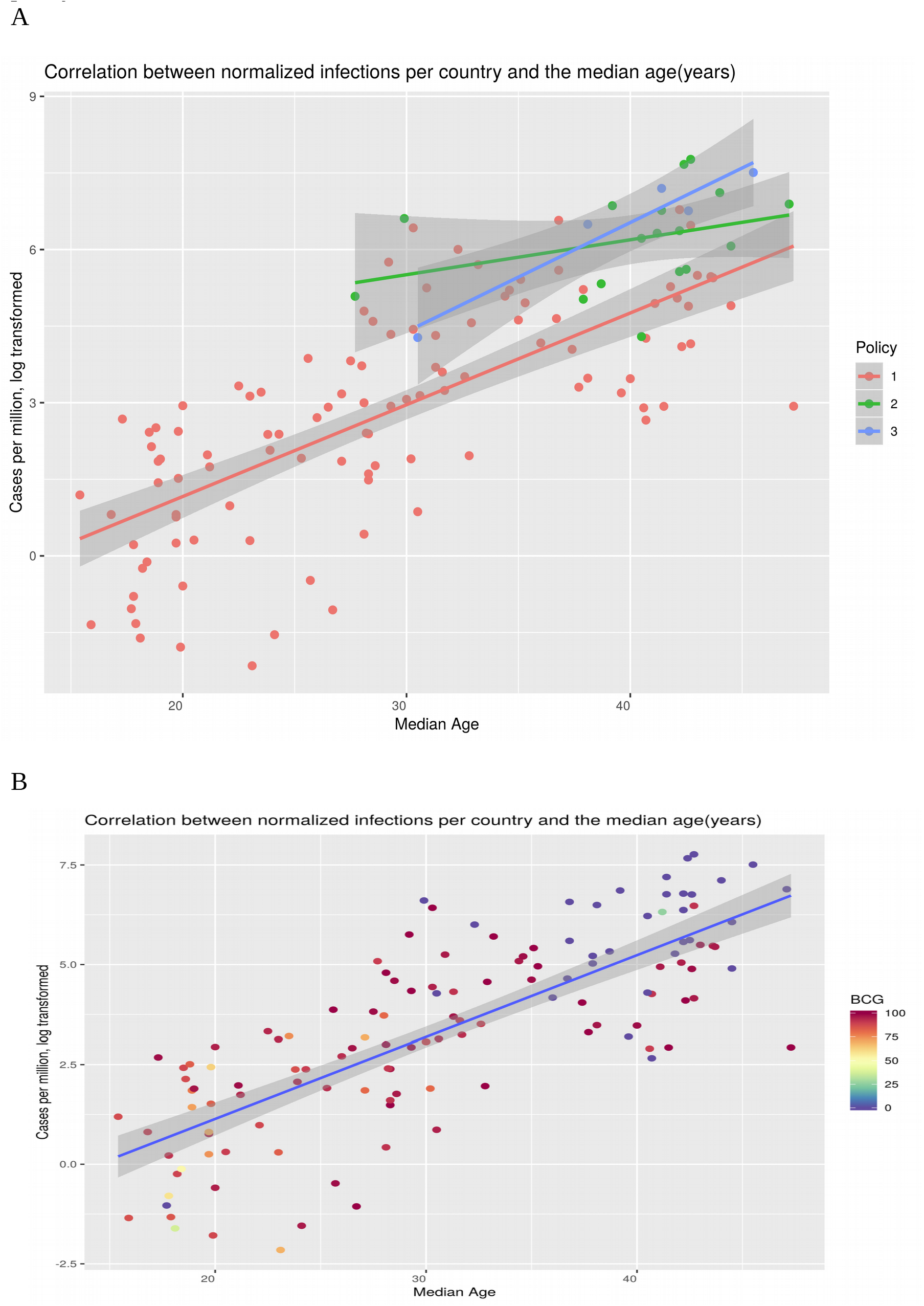
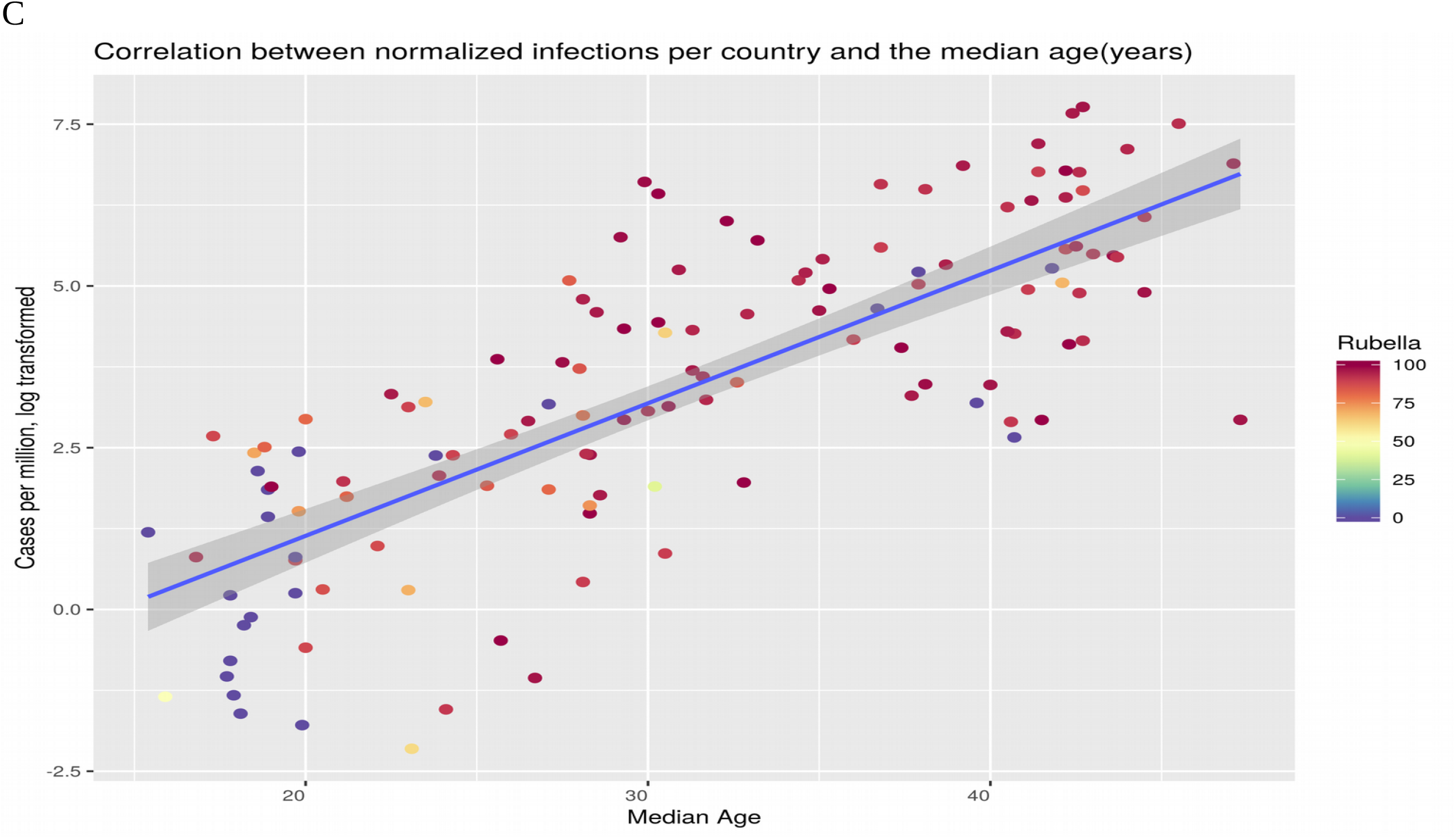
Relationship between median age per country and infection rates in the context of BCG policy

I also explored potential connection between countries with higher rubella immunization rates vs those with lower rates (separated in categories by 50% threshold) and COVID19 infections. While this variable on its own showed significant association (p<0.0001) with the observed infection rates per country, it appeared that the effect is the opposite of what would be expected (Figure 2C) with countries with low immunization rates scoring better in terms of infection rate. After the inclusion of other factors such as median age and income level this association was not significant at alpha=0.05 (p=0.056).

Since income levels are unlikely to drive infection rates I decided to compare the performance of median age and BCG policy. The data showed that median age explains the variance in the number of COVID19 cases better than the BCG policy either with or without income level adjustment (Figure 3). The median age explained 60% of the variability vs 30% for BCG policy. In a mixed model where income levels are considered a random factor median age again appears to be more important than BCG policy (Figure 4). BCG rates were again non-significant at p=0.0798.

**Figure 3.**
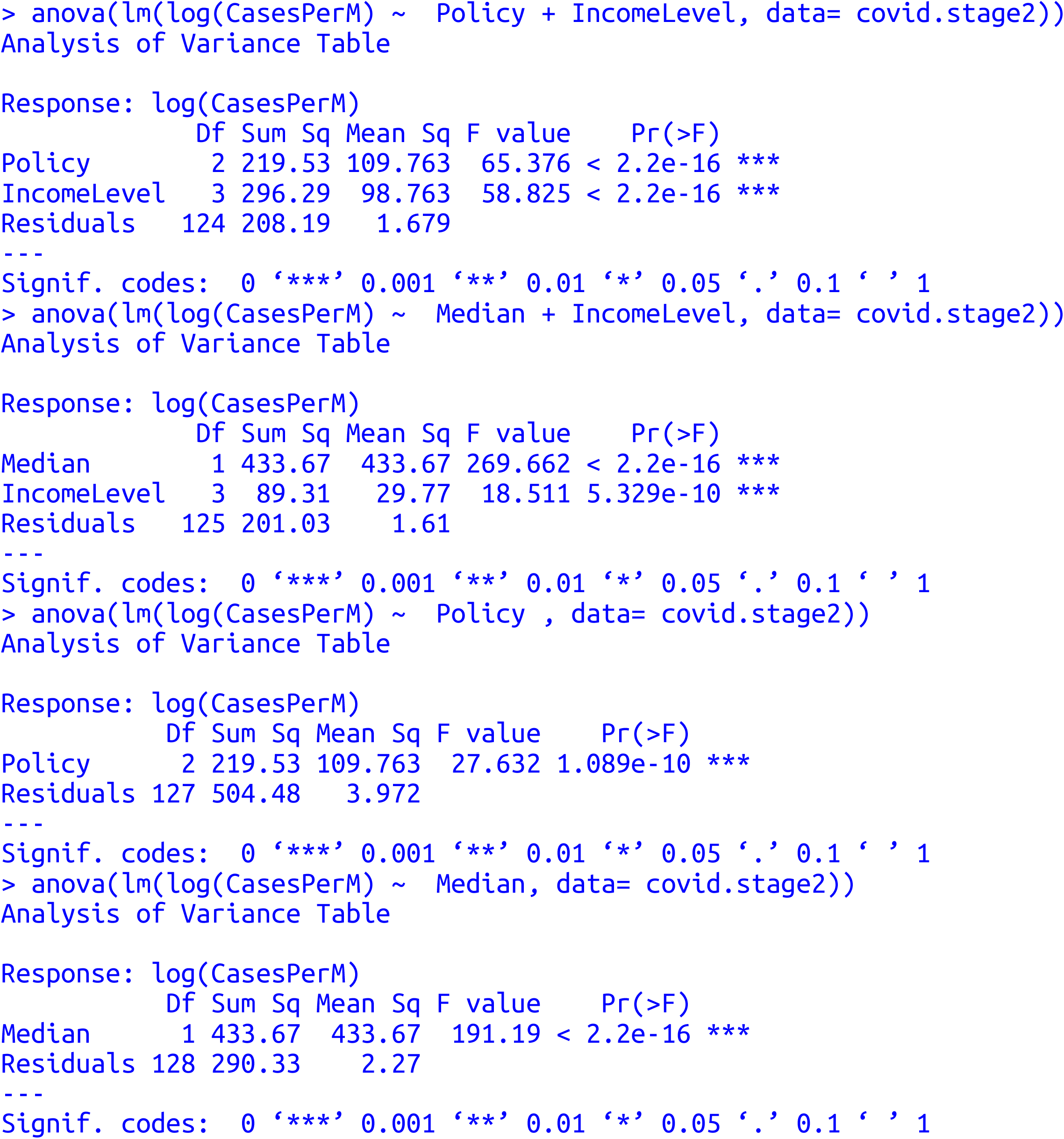
Effect of BCG policy or median age and infection rates in the presence and absence of income level effect

**Figure 4.**
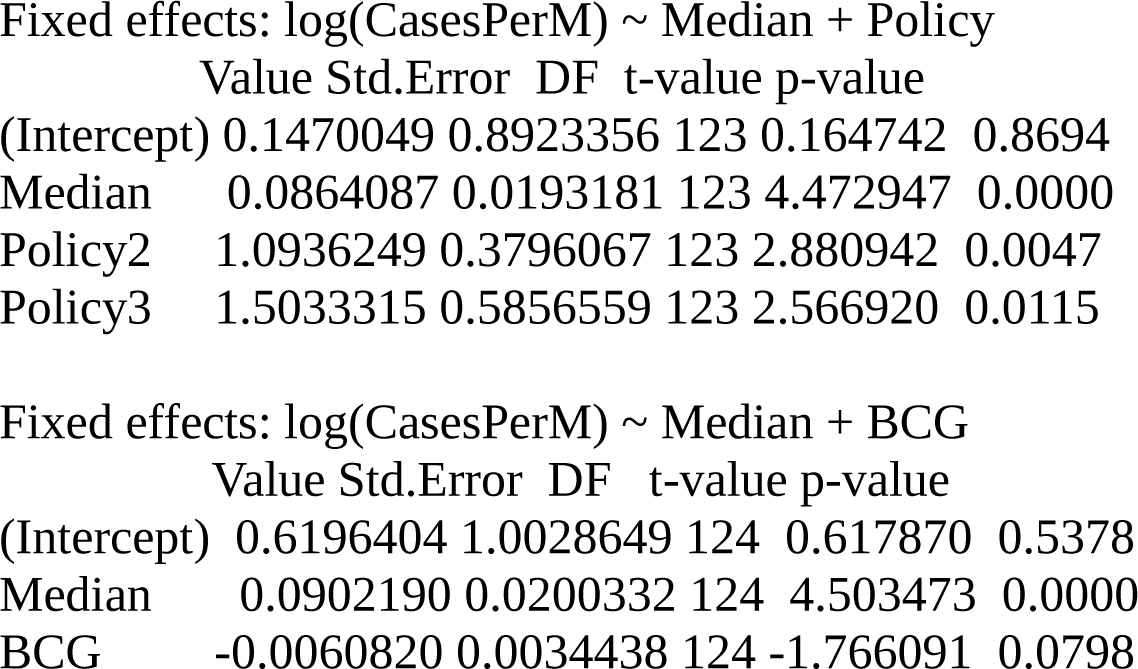
Mixed models used to evaluate associations between infection rates, median age, BCG policy and BCG immunization rates

Next, I looked at the median age distribution in different income levels and BCG policy categories (Figure 5). There was a strong association between median age and BCG policy with or without income level adjustment (p<0.0001). The same is true for median age and income level (p<0.0001). I also explored associations with mortality rates (Figure 6). Again, there was demonstrably better correlation between median age and mortality rates (R=0.653) compared to the correlation with start date of BCG vaccination policy reported in one of the studies (R=0.54)(4).

**Figure 5.**
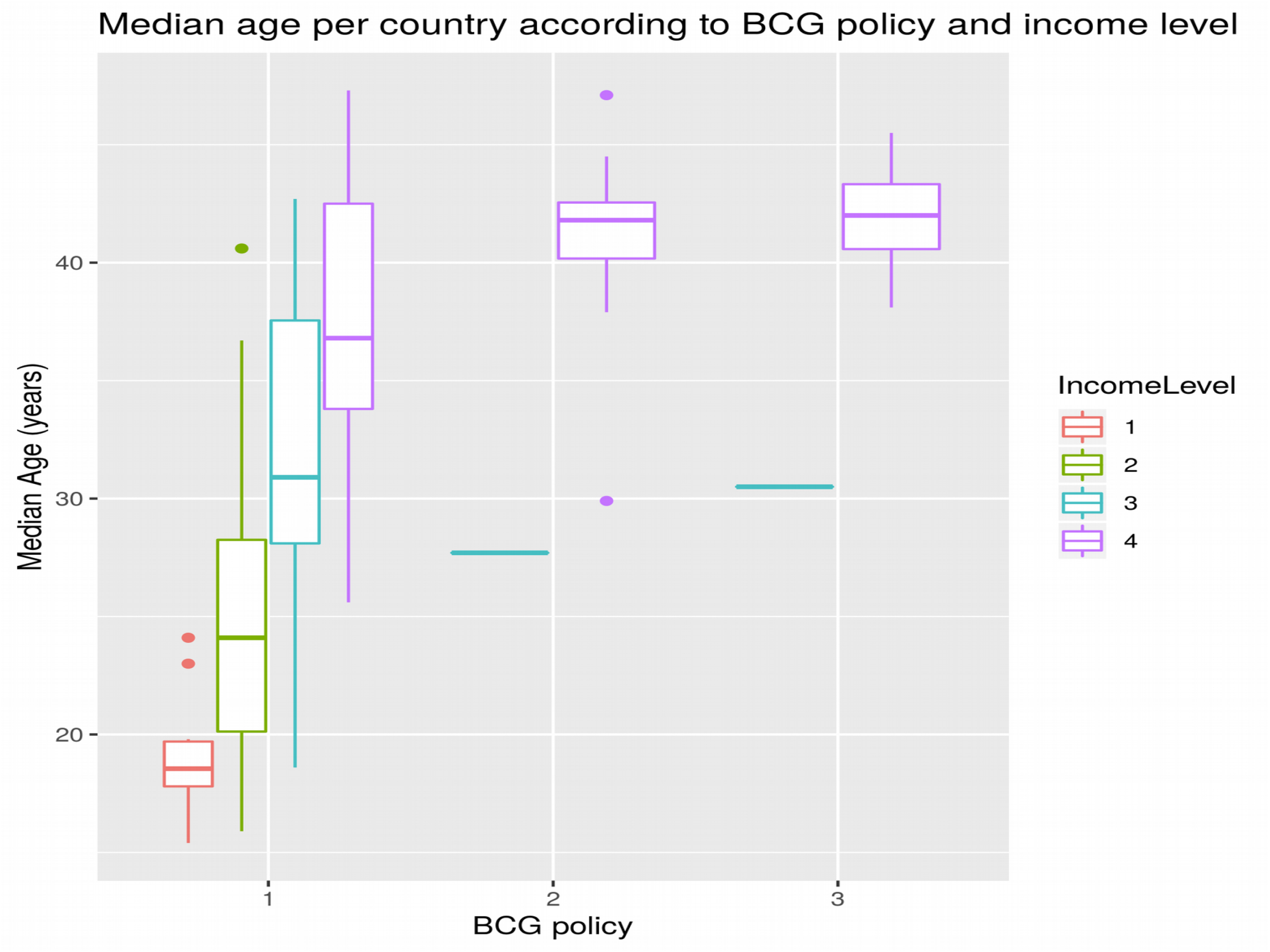
Relationship between median age and BCG policy in the context of income levels

**Figure 6.**
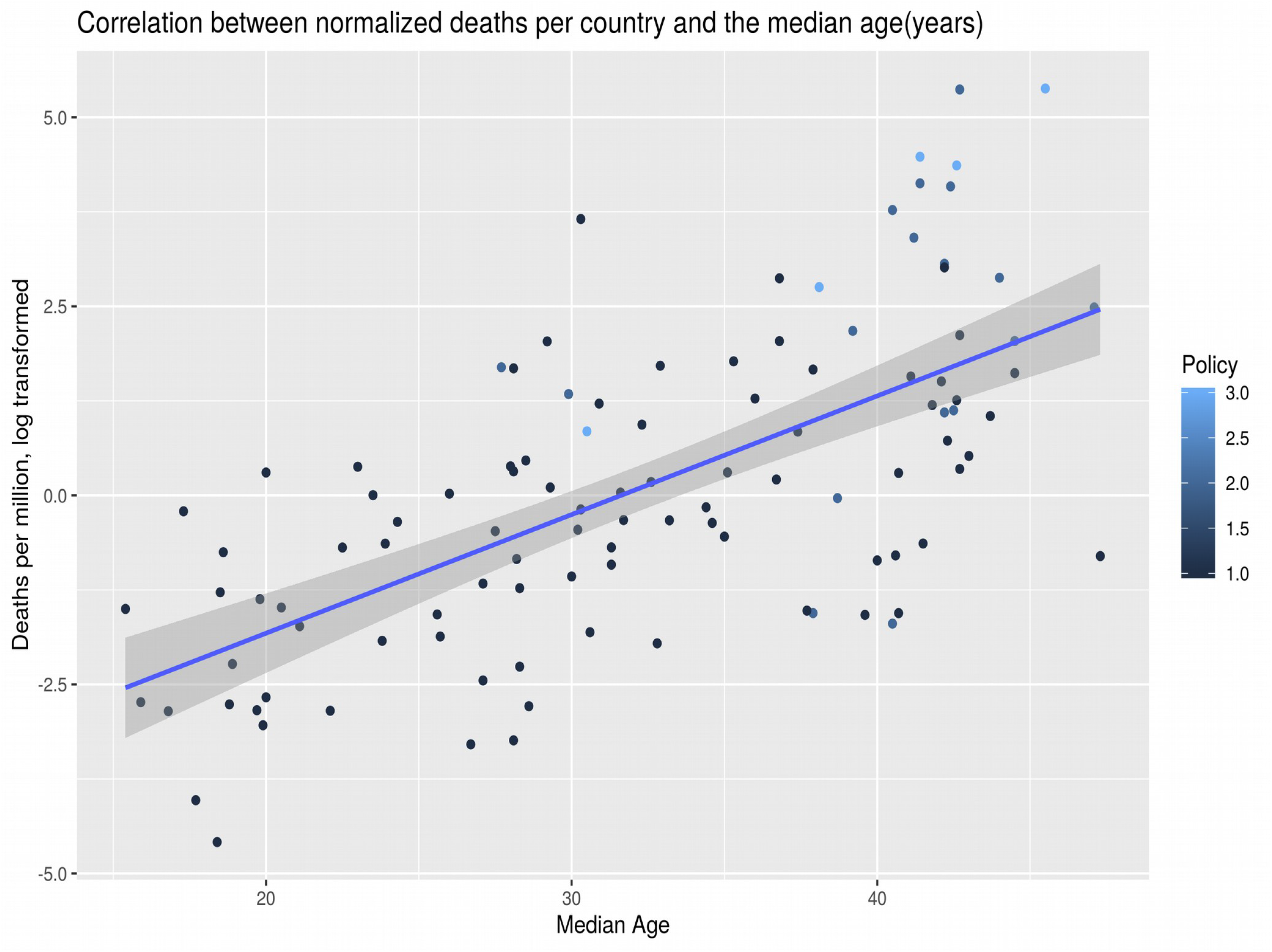

BMI was another strong confounding factor in the context of mortality rates(Figure 7). Countries with normal BMI were without exception in the policy category 1 (mandatory BCG vaccination). In addition, death rates were substantially higher in countries with high BMI (p<0.0001).

**Figure 7.**
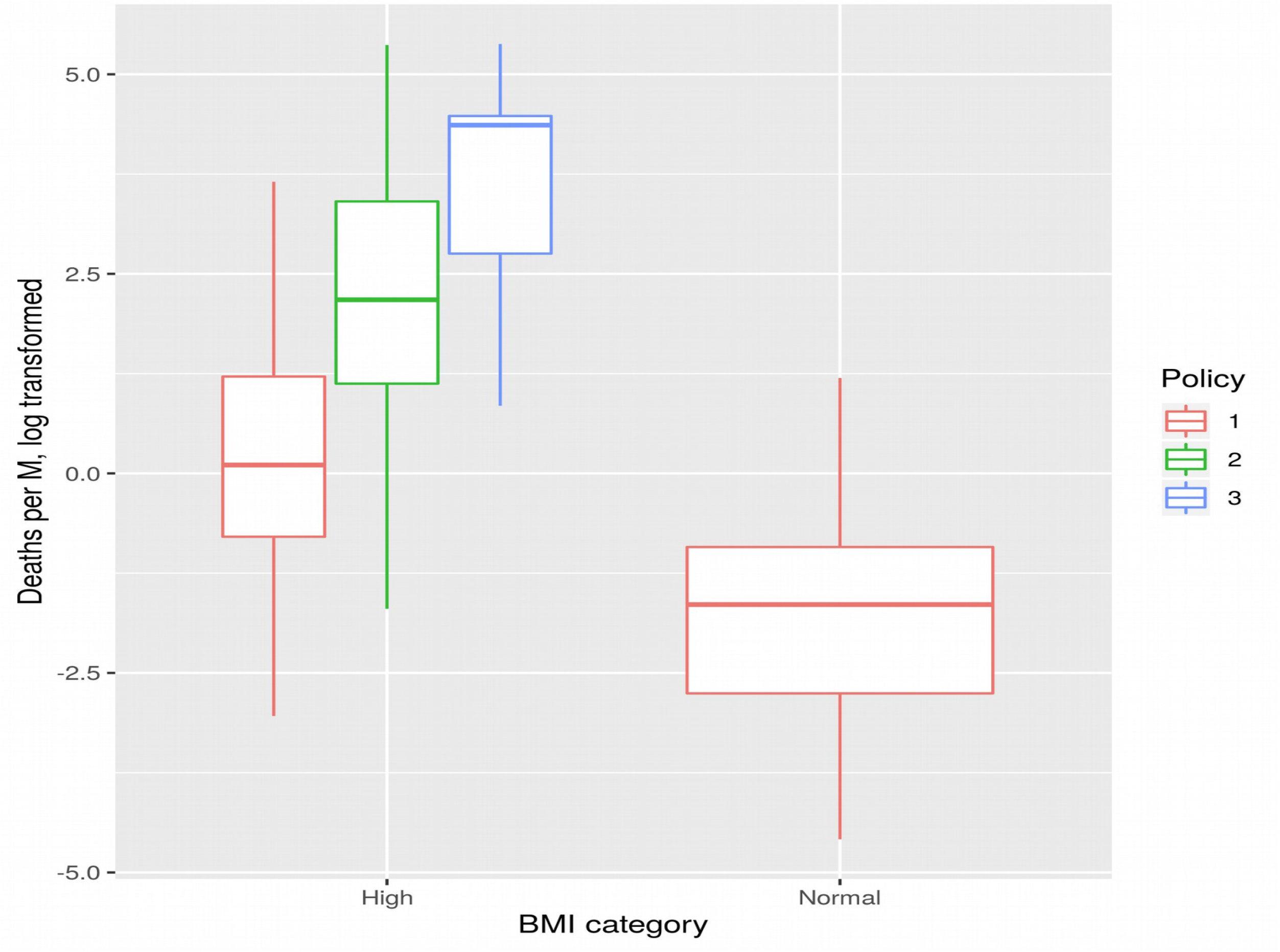
Association between normalized deaths from COVID19 and BMI (high>25) in the context of BCG policy

## Discussion

While observational studies are a valid and useful tool, there are also serious obstacles interpreting the data correctly(14). In the specific case of the correlation between BCG vaccination policy and COVID19 outcomes it is clear that important confounding factors may have been missed. An excellent outline of these obstacles was given by Emily MacLean(15). In addition to missing hidden factors, the critique in the blog goes further to challenge the biological plausibility of the BCG vaccine-COVID19 connection. It seems this is a very reasonable concern, given that the only established connection between BCG vaccination and protection against viral infections seems to be within the scope of actual anti-viral vaccines(2,3). On the other hand, the biological rational for causal link between age and COVID19 morbidity and mortality seems a lot more straightforward; if we follow the Occam’s razor we should prioritize this link over BCG vaccination. I also want to emphasize that the association observed in this work between infection rates and rubella immunization are almost certainly spurious. The arguments so far is that early childhood vaccinations might be protective, which is the opposite of our observation. Prior preclinical research(16) that was done during one of the previous coronavirus crisis shows clearly that childhood vaccinations are unlikely to drive different outcomes of COVID19 infections.

This is further enhanced by the data presented in this study. I need to emphasize that I have not included a number of other potentially confounding factors such as blood pressure, public policy (mandatory travel restrictions, use of masks, etc.) or time from first infection (start of community spread). Finally, any conclusive study will need to address the disagreement between BCG policy and actual BCG vaccination rates with the first still contributing to the regression model, whereas the second did not.

## Data Availability

All data is readily available from public sources. Processed data has been also made available.

https://github.com/kirovsa/covid19-bcg

## Note

The opinions expressed in this paper are personal and do not represent in any way Bristol Myers Squibb. No Bristol Myers Squibb resources were used to generate results or prepare this publication.

## Acknowledgment

I would like to thank Max Lau, PhD for critically reading this work and Kamen Kirov for editing the text.

